# An interpretable machine-learning model for predicting postoperative recovery quality after cardiovascular surgery: development, validation, and clinical applicability

**DOI:** 10.1101/2025.11.07.25339798

**Authors:** Luo Zhang, Bei Ma, Zhi Xing, Yudong Wang, Shunping Tian, Zhuan Zhang, Jianyou Zhang

**Author notes:** Corresponding Author: Jianyou Zhang Correspondence address: Department of Anesthesiology, the Affiliated Hospital of Yangzhou University, Yangzhou University, No. 368 Hanjiang Middle Road, Yangzhou 225012, China Corresponding author. Contributed equally to this work (Luo Zhang, Bei Ma, Zhi Xing).

## Abstract

**Objectives:** Quality of recovery (QoR) following cardiovascular surgery represents a key patient-centered outcome closely related to complications, hospital stay, and resource utilization. This study aimed to develop and validate an interpretable machine-learning model for predicting early postoperative recovery quality after cardiovascular surgery and to derive clinically actionable risk stratification to guide perioperative management.

**Methods:** We retrospectively analyzed 581 adult patients who underwent cardiovascular surgery at the Affiliated Hospital of Yangzhou University between March 2021 and September 2025. The primary endpoint was poor recovery, defined as QoR-15 < 90 on postoperative day 3. Predictor variables included demographic, ASA classification, emergency status, cardiopulmonary bypass (CPB), preoperative lactate, surgical duration, rebeating strategy, and modified Frailty Index (mFI). Data were randomly split 7:3 into training and test sets, with the final 20% of patients used for temporal external validation. Six ML algorithms include logistic regression (LR), K-nearest neighbors (KNN), Extremely Randomized Trees (ExtraTrees), Support Vector Machines (SVMs), Light Gradient Boosting Machine (LightGBM), and eXtreme Gradient Boosting (XGBoost) were compared using 10-fold cross-validation and hyperparameter optimization. Model discrimination, calibration, and clinical utility were evaluated using AUC, calibration plots, the Hosmer-Lemeshow test, and decision curve analysis (DCA). Model interpretability was assessed with SHapley Additive exPlanations (SHAP), and risk thresholds were derived from DCA for practical clinical stratification.

**Results:** Among the 581 patients, 173 (29.8%) experienced poor recovery. The XGBoost model achieved the best overall performance (AUC = 0.982, accuracy = 0.974, Hosmer–Lemeshow p = 0.791) with excellent calibration and temporal validation (AUC = 0.997). SHAP analysis identified five key predictors of poor recovery: female sex, higher ASA grade, elevated preoperative lactate (>2 mmol/L), longer operative duration, and greater frailty (mFI ≥ 0.25). Risk thresholds derived from DCA defined three clinical tiers-low (<0.15), intermediate (0.15-0.40), and high (>0.40)-for tailored postoperative management.

**Conclusions:** An interpretable XGBoost model accurately predicted postoperative recovery quality after cardiovascular surgery using routinely collected clinical data. The model’s transparency enables identification of modifiable risk factors and supports personalized perioperative optimization. Multicenter prospective validation and integration into perioperative decision-support systems are warranted to enhance recovery-oriented, patient-centered outcomes.

## Introduction

The quality of postoperative recovery (QoR) is a critical measure of patient-centered outcomes following surgery^1^. In cardiovascular and major vascular procedures, which involve substantial surgical trauma and complex perioperative management, accurately predicting and improving recovery remains a clinical challenge^2, 3^. Although continuous advancements in surgical and anesthetic techniques and perioperative care have led to a decline in in-hospital mortality rates for cardiac surgery, from 3.7% to 2.7% over the past decade^4^, recovery quality in these patients still lags behind other surgical populations.

Several patient and procedure-related factors, including impaired preoperative cardiac function, frailty, and systemic hypoperfusion, are known to contribute to poor recovery and higher complication rates^5–8^. The Quality of Recovery-15 (QoR-15) scale, a validated, patient-reported outcome measure,has been widely used for assessing postoperative recovery^9^. It captures five dimensions of recovery: physical comfort, emotional state, physical independence, psychological support, and pain. Its brevity, reliability, and responsiveness make it an ideal tool for assessing recovery quality in clinical research and routine practice^10^.

However, As Enhanced Recovery After Surgery (ERAS) protocols become increasingly integrated into perioperative management for cardiac surgery^11^, traditional risk assessment models have demonstrated limitations in identifying patients at risk for suboptimal recovery, few models have integrated these parameters into a unified predictive framework^12^.

To bridge this gap, the present study aimed to develop and validate an interpretable machine learning model that integrates comprehensive perioperative data to predict postoperative recovery quality after cardiovascular surgery. By identifying the most influential predictors and quantifying their contributions using SHapley Additive exPlanations (SHAP) values, we sought to provide an interpretable, data-driven framework that could support individualized perioperative management and early intervention.

## Methods

### Study design and ethical approval

This retrospective, observational study analyzed adult patients who underwent cardiovascular surgery at the Affiliated Hospital of Yangzhou University between March 2021 and September 2025. Clinical data were retrieved from the institutional electronic medical record (EMR) system under a standardized data extraction protocol. The study was approved by the institutional ethics committee (No. 2023YKL0109), which waived the requirement for informed consent owing to its retrospective nature. All procedures conformed to the principles of the Declaration of Helsinki.

### Participants

Inclusion criteria were: (1) age ≥18 years; (2) Completion of postoperative follow-up and QoR-15 assessment; (3) Availability of comprehensive clinical data, including demographic information, clinical characteristics, and laboratory results.

Exclusion criteria included: (1) Preoperative presence of severe cognitive impairment or psychiatric disorders that would not cooperate with the QoR-15 score; (2) Death or withdrawal of treatment within 24 hours after surgery.

### Outcome definition

The primary outcome was early postoperative recovery, measured by QoR-15 on postoperative day 3. QoR-15 questionnaire was used to assess postoperative recovery, consisting of 15 items across five domains: physical comfort (5 items), emotional state (4 items), physical independence (2 items), psychological support (2 items), and pain (2 items). The total score ranges from 0 to 150, with higher scores indicating better quality of recovery. A score of ≥ 90 was defined as good recovery, while scores < 90 indicated fair to poor recovery^13^. This threshold aligns with prior studies linking lower QoR-15 scores to prolonged hospitalization and higher complication risk.

### Data Collection

Clinical data were systematically extracted from the electronic medical record system using a standardized data collection protocol. To ensure data quality and consistency, all variables were independently reviewed by two trained researchers, with discrepancies resolved through consultation with a senior clinician.

Relevant patient information was retrieved from the electronic medical record system and categorized into general, surgical, and postoperative data. General data included gender, age, duration of postoperative follow-up, American Society of Anesthesiologists (ASA) physical status classification, preoperative lactate levels, comorbidities, emergency surgery status, and the use of cardiopulmonary bypass (CPB) during surgery. Surgical data encompassed the type and the duration of surgery, the duration of the operation, CPB time, aortic cross-clamp time, reperfusion status, intraoperative fluid therapy, and rewarming temperature. Postoperative data included length of stay in the Intensive Care Unit (ICU) and total hospital stay.

The Modified Frailty Index (mFI)^14^, a validated composite score based on 11 variables in the Canadian Study of Health and Aging, was used to classify patients as frail (mFI ≥ 0.25) or non-frail (mFI < 0.25).

### Data Preprocessing

Data preprocessing followed TRIPOD-ML recommendations to ensure reproducibility and transparency. Missing values were handled using multiple imputation techniques; for categorical variables, mode imputation was applied; for continuous variables, values were randomly selected near the mean within the range of standard deviation. The imputation error was controlled within 3% by comparing key statistical indicators (mean, median, and standard deviation) before and after imputation.

### Predictive Variables

Predictor variables were systematically selected based on the clinical relevance and biological plausibility and previous literature evidence. All laboratory indicators were measured using standardized protocols on the day of surgery or the following morning. The endpoint indicator of this study was poor postoperative recovery quality (QoR-15 < 90). Patients were categorized into poor recovery (score = 0) and good recovery (score = 1) groups based on their QoR-15 assessment completed within 72 hours after operation.

### Establishment of the Clinical Prediction Model

In this study, we adopted a multi-stage modeling approach to develop a predictive model for postoperative recovery quality. The analytical design combined conventional statistical techniques with advanced machine learning algorithms to achieve both reliable predictor selection and optimal model performance. Initially, univariate logistic regression was applied to identify potential factors associated with poor postoperative recovery. Variables showing statistical significance (*P* < 0.05) were subsequently included in the multivariate analysis. A stepwise forward selection procedure, guided by the likelihood ratio test, was then used to determine the independent effects of each variable while accounting for possible confounders. Ultimately, the final model incorporated only those predictors that were both statistically significant (*P* < 0.05) and clinically meaningful.

To enhance predictive accuracy and explore complex non-linear relationships among variables, we integrated six algorithms such as the logistic regression (LR), support vector machine (SVM), K-nearest neighbors (KNN), Extremely randomized trees (Extra Trees), eXtreme Gradient Boosting (XGBoost), and Light Gradient Boosting Machine (LightGBM) to develop the composite model. Specifically, the SVM model utilized multiple kernel functions to determine optimal decision boundaries in high-dimensional feature spaces, making it particularly effective in detecting complex, non-linear associations among predictors. The KNN algorithm, as a non-parametric and intuitive approach, classified samples according to their similarity with neighboring data points, thereby serving as a straightforward yet interpretable benchmark for distance-based prediction. Meanwhile, both XGBoost and LightGBM incorporated L1/L2 regularization to reduce overfitting and employed adaptive strategies to manage missing values, enhancing performance on large and heterogeneous datasets. Collectively, this integrative modeling strategy allowed for a more stable and accurate identification of key predictive variables.

Data were randomly stratified into training (70%) and internal test (30%) sets, ensuring balanced class distributions. A fixed random seed was used for reproducibility. The split ratio followed empirical practice for clinical prediction modeling, balancing learning stability with adequate validation sample size.

Model generalization was examined by repeated 10-fold cross-validation and permutation testing (n=1000). Differences in AUC between cross-validation and test sets <0.02 were considered evidence of minimal overfitting.

The final model (XGBoost) incorporated the most stable and clinically meaningful predictors. Feature contribution was quantified using SHapley Additive exPlanations (SHAP), enabling visualization of each variable’s marginal impact on model output and supporting clinical interpretability.

### Temporal external validation

To evaluate temporal robustness, an independent cohort comprising the final 20% of patients (n=116) who underwent surgery later in the study period was reserved for temporal external validation. All preprocessing and hyperparameters were applied unchanged; no model retraining occurred. This procedure assessed real-world stability across time within the same clinical environment.

Future multicenter studies are planned to confirm model generalizability across different institutions and patient populations.

### Model Performance Evaluation

The predictive performance of each model was comprehensively assessed using a series of evaluation metrics. The area under the receiver operating characteristic curve (AUC) was used to measure the ability of the model to discriminate between patients with good and poor postoperative recovery. Accuracy (Acc) reflected the overall proportion of correctly classified cases, providing a general measure of predictive reliability; Sensitivity (Sn) indicated the model’s capacity to correctly identify patients with poor recovery outcomes, whereas specificity (Sp) represented its ability to recognize those with good recovery; The positive predictive value (PPV) denoted the likelihood that patients predicted to have poor recovery truly experienced it, while the negative predictive value (NPV) described the probability that patients predicted to recover well actually did so.

### Statistical Analysis

All statistical analyses were performed using SPSS version 26.0 (IBM Corp., Armonk, NY, USA), Python 3.10 (scikit-learn package) and MATLAB R2024a (The MathWorks, Inc., Natick, MA, USA). XGBoost, LightGBM, and SHAP packages for machine learning implementation. Continuous variables were tested for normality using the Kolmogorov-Smirnov test. Normally distributed data were presented as mean ± standard deviation (SD) and compared using independent-sample t-tests, non-normally distributed data were expressed as median (interquartile range, IQR). Categorical variables were expressed as frequencies and percentages and compared using the Chi-square test or Fisher’s exact test. Calibration curves, the Hosmer-Lemeshow test, and decision curve analysis (DCA) were used to evaluate calibration and clinical utility. Statistical significance was defined as *P* < 0.05 (two-sided).

### Reporting guideline compliance

This study adheres to the Transparent Reporting of a multivariable prediction model for Individual Prognosis Or Diagnosis (TRIPOD-ML) extension for machine-learning-based models and the RECORD (REporting of studies Conducted using Observational Routinely-collected health Data) guidelines. All analyses and reporting procedures were conducted in accordance with these standards to ensure methodological transparency, reproducibility, and interpretability.

## Results

### Baseline Characteristics

The flow chart of this study is shown in Fig. 1. A total of 581 patients undergoing cardiovascular surgery were included in the analysis, among which 173 patients had poor recovery. As shown in Table 1, multiple factors were identified as significantly associated with QoR-15 in patients. These included gender(P = 0.047), age(*P* < 0.001), ASA classification(*P* < 0.001), preoperative lactate level(*P* < 0.001), emergency(*P* < 0.001), cardiopulmonary bypass(*P* = 0.016), surgery duration(*P* < 0.001) rebeating strategies(*P* = 0.015), duration of ICU stay(*P* < 0.001), total hospitalization days(*P* < 0.001) and the mFI (*P* = 0.003). Multivariable logistic regression analysis was performed using variables that were statistically significant in the univariate analysis. As shown in Table 2, five factors were independently associated with excellent postoperative recovery. These included gender (OR = 1.70, 95% CI: 1.09-2.66, *P* = 0.02), indicating that male patients were significantly more likely to achieve excellent recovery; ASA classification (OR = 2.5, 95% CI: 1.45-4.31, *P* = 0.001) and preoperative lactate level (OR = 3.2, 95% CI: 1.67-6.15, *P* = 0.001) were both strong predictors of recovery quality, with higher ASA grades and elevated lactate levels associated with poorer outcomes; The duration of the surgery(OR = 1.01, 95% CI: 1.00-1.02, *P* = 0.001) was oppositively associated with QoR-15 scores, suggesting improved recovery quality with shorten the operation time. In addition, mFI was a significant predictor (OR = 2.20, 95% CI: 1.04-4.68, *P* = 0.04), with frail patients showing significantly lower odds of excellent recovery. In contrast, other variables did not show statistically significant associations with recovery quality in the

**Figure 1.**
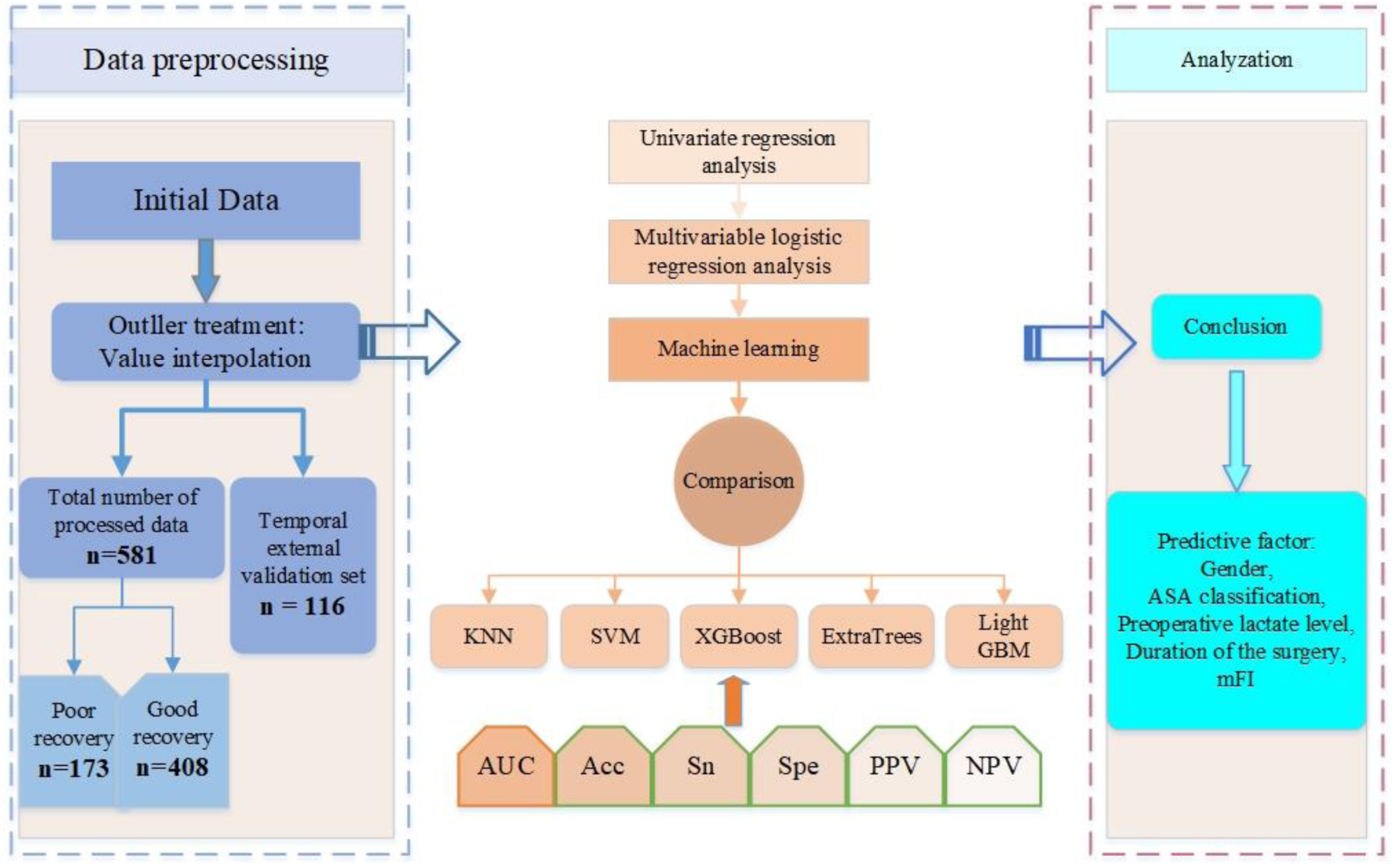
Flowchart of this study

**Table 1.**
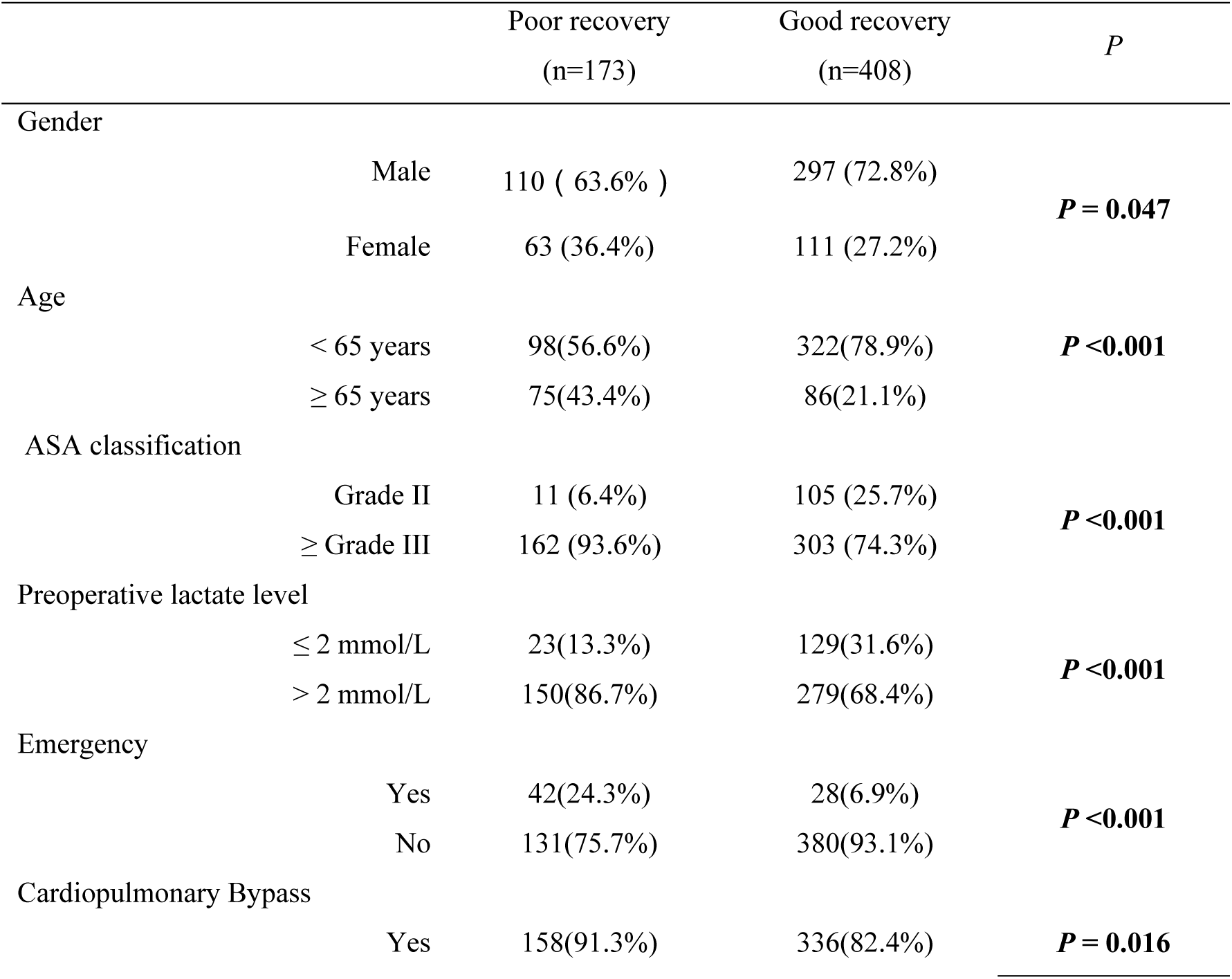
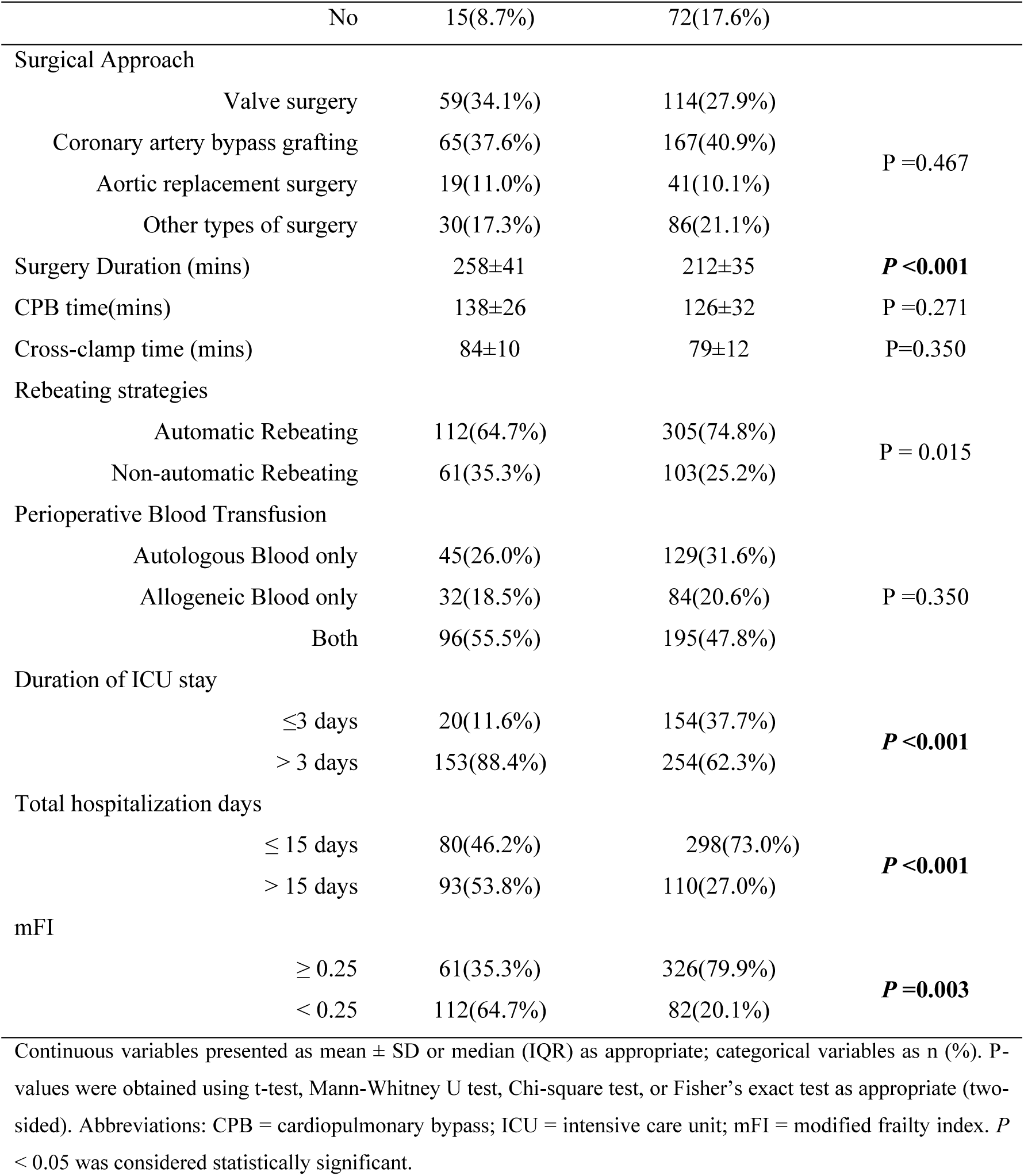
Univariate analysis of factors influencing patient recovery quality.

**Table 2.** Factors affecting patient recovery quality: multivariate analysis.

|  | Variable coefficient | Standard error | Wald | *P* | OR | 95%CI |  |
| --- | --- | --- | --- | --- | --- | --- | --- |
|  |  |  |  |  |  | Upper limit | Lower limit |
| Gender(Female vs. Male) | 0.53 | 0.23 | 5.43 | **0.020** | 1.70 | 1.09 | 2.66 |
| ASA classification | 0.92 | 0.28 | 10.83 | **0.001** | 2.50 | 1.45 | 4.31 |
| Preoperative lactate level | 1.16 | 0.33 | 12.18 | **0.001** | 3.20 | 1.67 | 6.15 |
| Surgery Duration | 0.30 | 0.02 | 8.76 | **0.001** | 1.35 | 1.12 | 1.62 |
| mFI | 0.79 | 0.38 | 4.20 | **0.040** | 2.20 | 1.04 | 4.68 |

OR, odds ratio; CI, confidence interval, $P < 0.05$ was considered statistically significant.

### Model Performance

The reliability of the constructed model was validated in a rigorously designed cohort. Notably, no significant gender differences were observed between poor recovery and good recovery patients, suggesting that potential confounding effects related to sex were effectively balanced across groups. Using a comprehensive multimodal dataset encompassing baseline characteristics, laboratory findings, and other perioperative datas, the predictive performance of six algorithms: LR, Extra Trees, LightGBM, SVM, XGBoost, and KNN was systematically compared within an integrated learning framework. We observed that the XGBoost algorithm showed optimal overall performance in the independent validation set: as shown in Table 3, the test *P* values of XGBoost in the training and test datasets are 0.997 and 0.791 (P > 0.05), respectively, indicating that its predicted probability is highly consistent with the actual event distribution and has excellent calibration performance. The integrated calibration curve indicated that the predicted values were in good agreement with the observed values between the training and testing cohorts of the XGBoost algorithm. These data suggested that the XGBoost model exhibited optimal overall performance in the training and test cohorts, and the calibration curves in both cohorts is shown in Fig. 2a, b. The area under the receiver operating characteristic curve of the working characteristics of the subjects was 0.982 (95% confidence interval (CI) 0.974 - 1.000) (Fig. 2c, d), the Acc was 0.974 (95% CI 0.964 - 0.984), the Sn and Sp were 0.962 and 0.980, respectively, while the positive and negative predictive values were 0.962 and 0.980, respectively see Table 4 for details. The decision curve analysis (DCA) was adopted to assess each model. As shown in Fig. 2e, f, DCA revealed that XGBoost had the best performance, with a significantly higher net benefit compared with the other methods in the range of medium and high-risk thresholds. This confirmed that XGBoost could effectively integrate non-lin-ear associations between copper metabolic abnormalities and organ function indicators when dealing with multidimensional medical data, providing a high-precision tool for early warning detection of poor recovery of patients undergoing cardiovascular surgery. Furthermore, to verify the model’s general applicability, external validation was conducted in this study. This demonstrated that XGBoost still performed optimally, with an area under the curve (AUC) of 0.997 (95% CI 0.994-1.000), an Acc of 0.974, a Sen of 0.962 and a Spe of 0.980, as well as a PPV and NPV of 0.962 and 0.980(Fig. 3 and Table 5). The final XGBoost model achieved excellent discrimination in the temporally independent external validation cohort, confirming its generalizability over time.

**Table 3.** Hosmer_lemeshow_test in the training and test dataset.

| Model | Train P value | Test P value |
| --- | --- | --- |
| LR | 0.867 | 0.739 |
| KNN | 0.277 | 0.011 |
| ExtraTrees | 0.235 | 0.114 |
| XGBoost | 0.997 | 0.791 |
| LightGBM | 0.188 | 0.156 |
| rbf_SVM | 0.981 | 0.432 |
| linear_SVM | 0.998 | 0.059 |
| sigmoid_SVM | 0.936 | 0.241 |
| poly_SVM | 0.653 | 0.527 |

Temporal external validation set (n = 116, 20% of the total cohort; temporally independent from development data). Used for model generalizability assessment.

**Figure 2.**
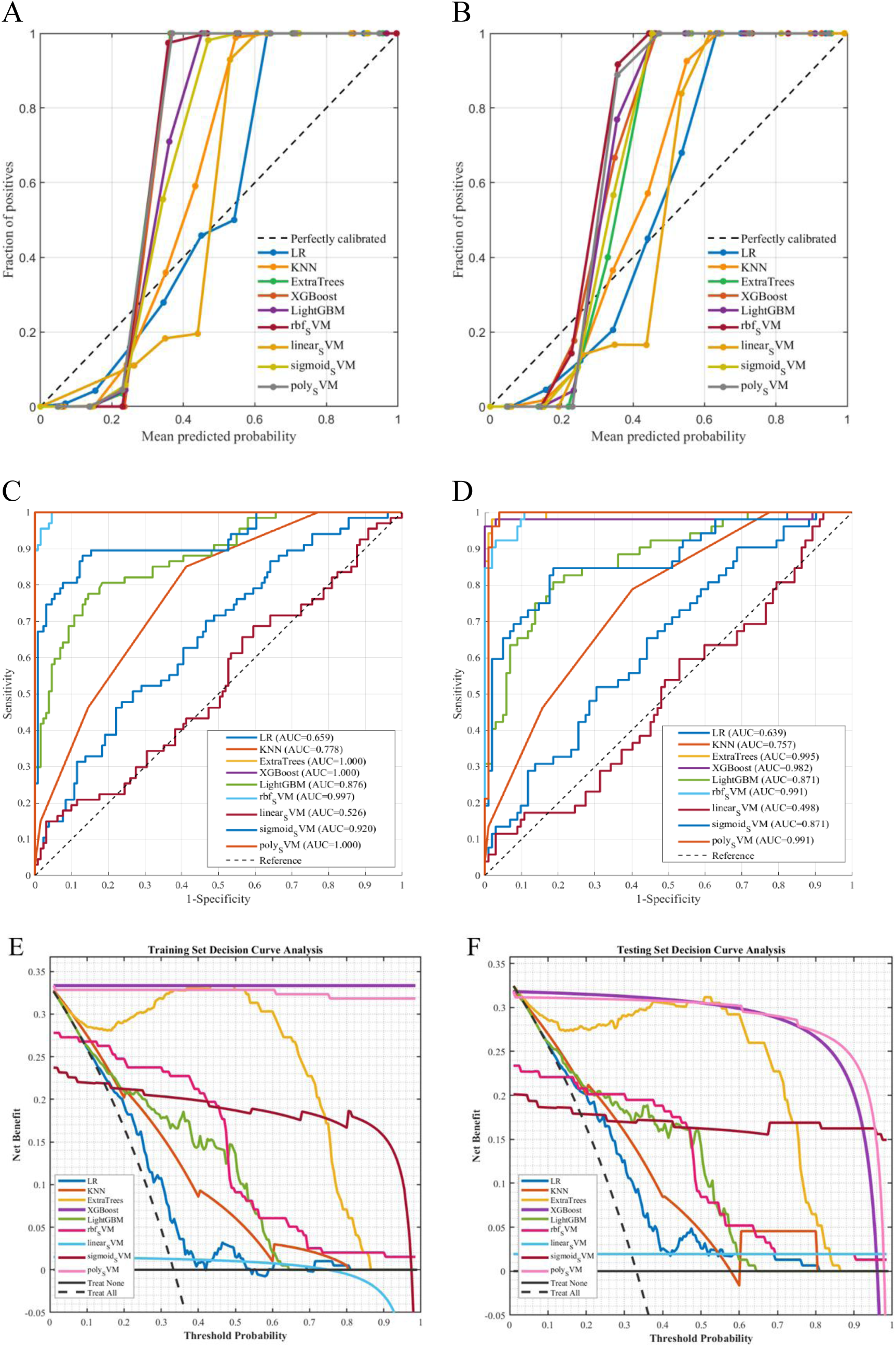
Model calibration curves, receiver operating characteristic (ROC) curves, and decision curve analysis (DCA) for the development and test cohorts. **Panels A-B**: calibration plots comparing predicted vs observed probability for XGBoost in training (A) and test (B) cohorts; dashed line = ideal calibration. **Panel C-D**: ROC curves (AUC reported). **Panel E-F**: Decision curve analysis (net benefit plotted against threshold probability). All AUCs and 95% confidence intervals were estimated using bootstrap resampling (1,000 replicates). XGBoost was applied to held-out cohorts without retraining. Abbreviations: AUC = area under the ROC curve; CI = confidence interval.

**Table 4.** Machine learning modeling analysis in the train and test dataset.

| Cohort | Model | AUC | AUC<br>95%CI | Acc | Acc<br>95%CI | Sen | Spe | PPV | NPV |
| --- | --- | --- | --- | --- | --- | --- | --- | --- | --- |
| Train | LR | 0.659 | 0.635,<br>0.793 | 0.667 | 0.648,<br>0.706 | 0.149 | 0.932 | 0.536 | 0.682 |
| Test | LR | 0.639 | 0.581,<br>0.711 | 0.662 | 0.616,<br>0.674 | 0.154 | 0.922 | 0.500 | 0.681 |
| Train | KNN | 0.778 | 0.748,<br>0.780 | 0.722 | 0.702,<br>0.777 | 0.463 | 0.855 | 0.620 | 0.757 |
| Test | KNN | 0.757 | 0.706,<br>0.776 | 0.714 | 0.693,<br>0.774 | 0.462 | 0.843 | 0.600 | 0.754 |
| Train | Extra Trees | 1.000 | 1.000,<br>1.000 | 0.995 | 0.991,<br>0.999 | 0.985 | 1.000 | 1.000 | 0.992 |
| Test | Extra Trees | 0.995 | 0.988,<br>0.998 | 0.967 | 0.949,<br>0.973 | 0.942 | 0.981 | 0.961 | 0.971 |
| Train | XG Boost | 1.000 | 1.000,<br>1.000 | 1.000 | 1.000,<br>1.000 | 1.000 | 0.989 | 0.971 | 0.970 |
| Test | XG Boost | 0.982 | 0.974,<br>1.000 | 0.974 | 0.964,<br>0.984 | 0.962 | 0.980 | 0.962 | 0.980 |
| Train | Light GBM | 0.876 | 0.857,<br>0.907 | 0.778 | 0.678,<br>0.828 | 0.373 | 0.985 | 0.926 | 0.754 |
| Test | Light GBM | 0.871 | 0.802,<br>0.885 | 0.773 | 0.761,<br>0.781 | 0.365 | 0.981 | 0.905 | 0.752 |
| Train | rbf SVM | 0.998 | 0.995,<br>1.004 | 0.949 | 0.942,<br>0.968 | 0.851 | 0.999 | 0.988 | 0.929 |
| Test | rbf SVM | 0.991 | 0.959,<br>0.998 | 0.929 | 0.902,<br>0.944 | 0.788 | 0.999 | 0.998 | 0.903 |
| Train | linear SVM | 0.526 | 0.483,<br>0.545 | 0.672 | 0.642,<br>0.690 | 0.445 | 0.992 | 0.750 | 0.671 |
| Test | linear SVM | 0.498 | 0.433,<br>0.556 | 0.675 | 0.633,<br>0.691 | 0.385 | 0.990 | 0.732 | 0.670 |
| Train | sigmoid SVM | 0.920 | 0.892,<br>0.928 | 0.879 | 0.865,<br>0.893 | 0.776 | 0.932 | 0.852 | 0.891 |
| Test | sigmoid SVM | 0.871 | 0.857,<br>0.913 | 0.831 | 0.814,<br>0.891 | 0.692 | 0.902 | 0.783 | 0.852 |
| Train | poly SVM | 1.000 | 1.000,<br>1.000 | 0.995 | 0.991,<br>0.999 | 0.985 | 1.000 | 1.000 | 0.992 |
| Test | poly SVM | 0.991 | 0.946,<br>1.070 | 0.961 | 0.864,<br>0.988 | 0.923 | 0.980 | 0.961 | 0.962 |

Metrics reported: AUC (95% CI by bootstrap, 1,000 replicates), accuracy (Acc), sensitivity (Sen), specificity (Spe), positive predictive value (PPV), negative predictive value (NPV). The XGBoost model reported in the table is the final model trained on the development set and applied to test/temporal cohorts without retraining. Abbreviations: CI = confidence interval; AUC = area under ROC curve.

**Figure 3.**
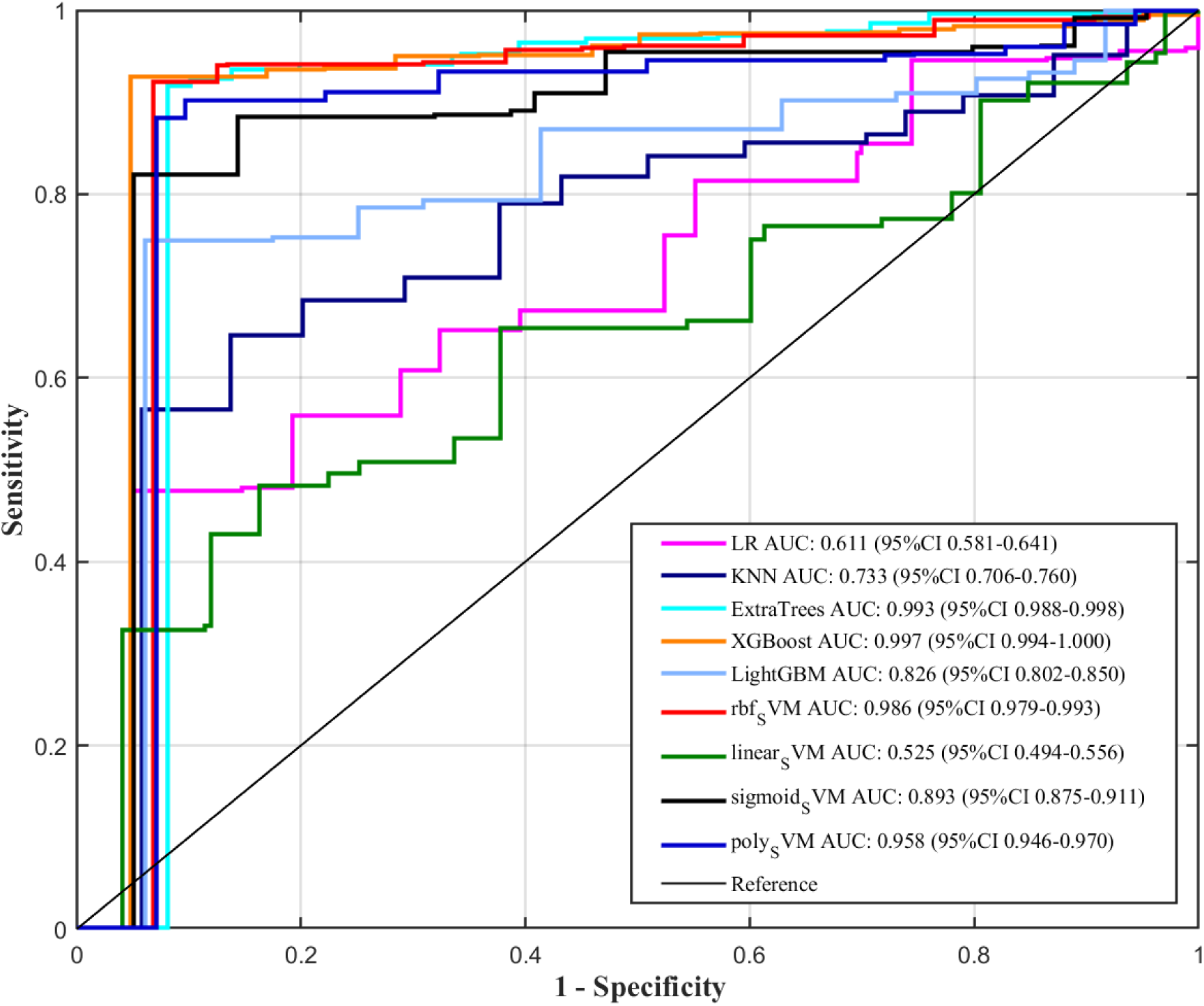
ROC curve of XGBoost applied to the temporal validation cohort (last 20% of patients). Model was frozen (no refitting) and performance evaluated on n =116 independent temporal patients. AUC and 95% CI obtained from 1,000 bootstrap replications.

**Table 5.**
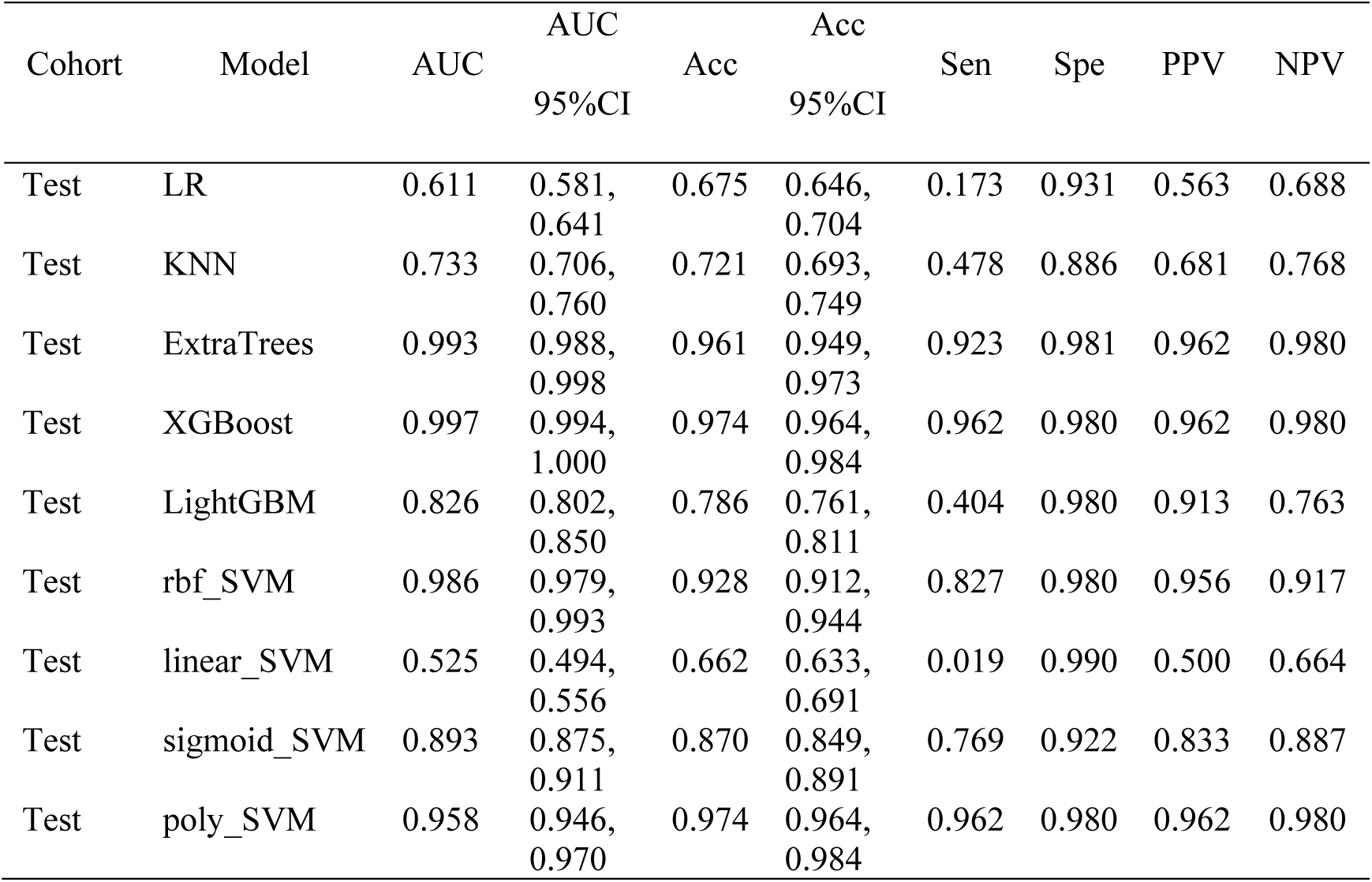
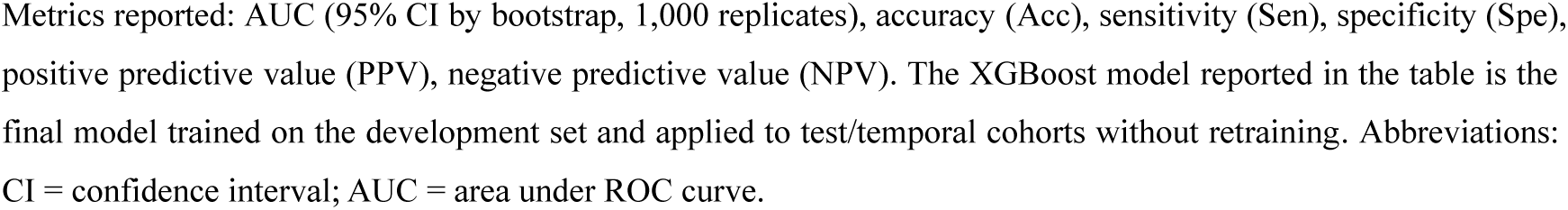
Machine learning modeling analysis in the test dataset for external validation.

| Cohort | Model | AUC |  | Acc |  | Sen | Spe | PPV | NPV |
| --- | --- | --- | --- | --- | --- | --- | --- | --- | --- |
|  |  | AUC | 95%CI | Acc | 95%CI |  |  |  |  |
| Test | LR | 0.611 | 0.581, 0.641 | 0.675 | 0.646, 0.704 | 0.173 | 0.931 | 0.563 | 0.688 |
| Test | KNN | 0.733 | 0.706, 0.760 | 0.721 | 0.693, 0.749 | 0.478 | 0.886 | 0.681 | 0.768 |
| Test | ExtraTrees | 0.993 | 0.988, 0.998 | 0.961 | 0.949, 0.973 | 0.923 | 0.981 | 0.962 | 0.980 |
| Test | XGBoost | 0.997 | 0.994, 1.000 | 0.974 | 0.964, 0.984 | 0.962 | 0.980 | 0.962 | 0.980 |
| Test | LightGBM | 0.826 | 0.802, 0.850 | 0.786 | 0.761, 0.811 | 0.404 | 0.980 | 0.913 | 0.763 |
| Test | rbf_SVM | 0.986 | 0.979, 0.993 | 0.928 | 0.912, 0.944 | 0.827 | 0.980 | 0.956 | 0.917 |
| Test | linear_SVM | 0.525 | 0.494, 0.556 | 0.662 | 0.633, 0.691 | 0.019 | 0.990 | 0.500 | 0.664 |
| Test | sigmoid_SVM | 0.893 | 0.875, 0.911 | 0.870 | 0.849, 0.891 | 0.769 | 0.922 | 0.833 | 0.887 |
| Test | poly_SVM | 0.958 | 0.946, 0.970 | 0.974 | 0.964, 0.984 | 0.962 | 0.980 | 0.962 | 0.980 |

Metrics reported: AUC (95% CI by bootstrap, 1,000 replicates), accuracy (Acc), sensitivity (Sen), specificity (Spe), positive predictive value (PPV), negative predictive value (NPV). The XGBoost model reported in the table is the final model trained on the development set and applied to test/temporal cohorts without retraining. Abbreviations: CI = confidence interval; AUC = area under ROC curve.

### Identification of Predictive Factors for the Postoperative Recovery Quality Prediction Model

Feature importance was analyzed using the SHapley Additive exPlanations (SHAP) framework to enhance model interpretability from both global and individual perspectives. As shown in Figure 4A, features with a wider distribution of SHAP values exerted a greater influence on the prediction outcomes. The mean absolute SHAP value for each variable was then calculated to quantify its overall contribution to the XGBoost classifier and displayed as a ranked bar plot in Figure 4B. This visualization intuitively demonstrated the relative importance of each predictor within the model. Consistent with the Gini impurity-based feature selection, the most influential predictors associated with postoperative recovery quality included female sex, higher ASA classification, elevated preoperative lactate levels, longer operative duration, and higher mFI. These results offer a robust interpretive foundation for the model and identify clinically relevant factors that may guide early perioperative interventions aimed at improving postoperative recovery outcomes.

**Fig. 4.**
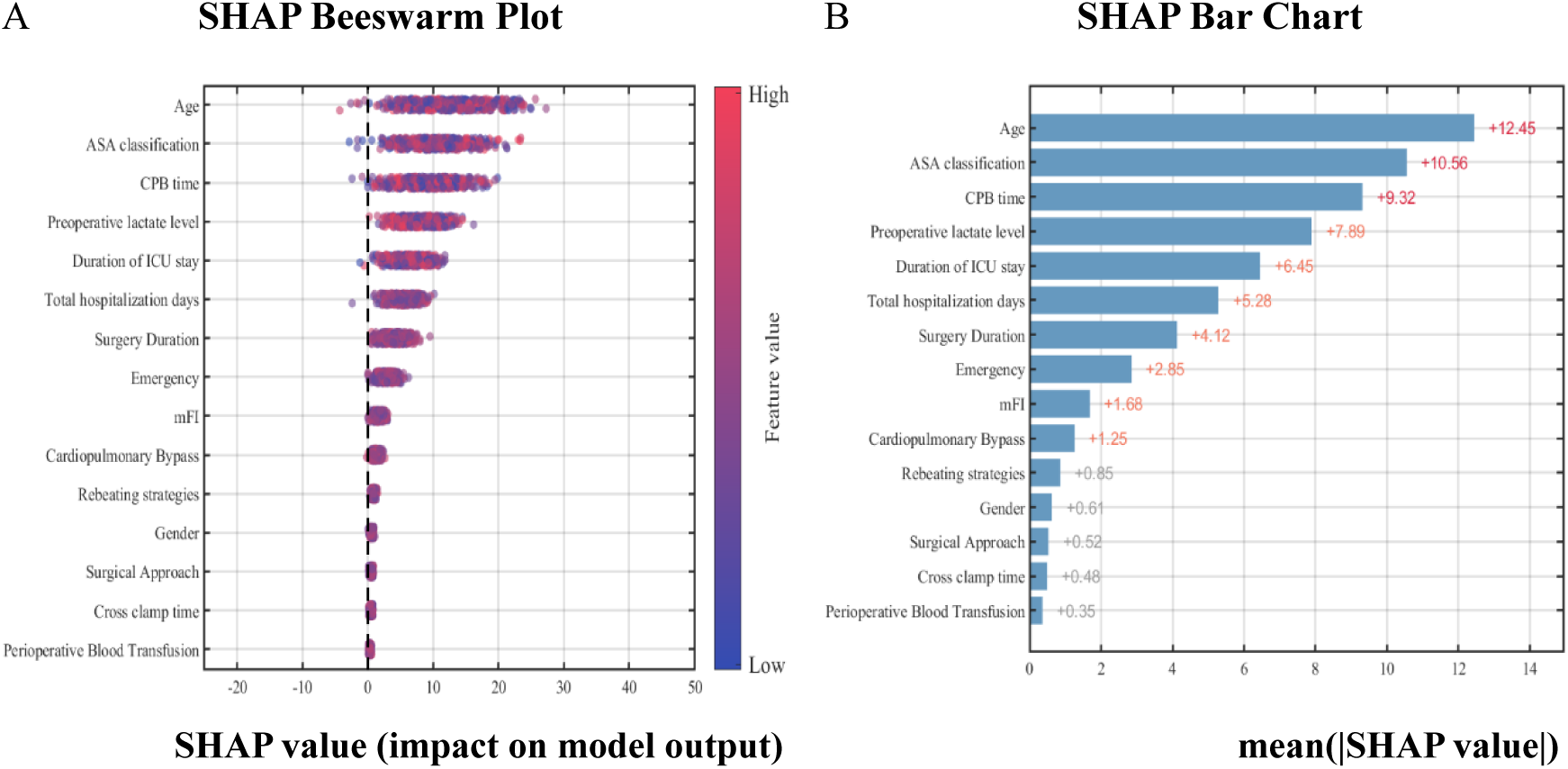
SHAP-based interpretability analysis for the XGBoost model. (A) Beeswarm plot showing per-sample SHAP values (horizontal axis indicates SHAP impact on model output; each point represents one patient). Color scale indicates original feature value (red = higher value; blue = lower value). (B) Mean absolute SHAP values ranking overall feature importance. Positive SHAP values increase predicted risk of poor recovery; negative values decrease predicted risk. Abbreviation: SHAP = SHapley Additive exPlanations.

## Discussion

In this retrospective study of 581 patients undergoing cardiovascular surgery, we developed and validated an interpretable machine learning (ML) model to predict early postoperative recovery quality as measured by the QoR-15 scale. Among six candidate algorithms, the XGBoost classifier achieved the best discrimination (AUC 0.982) and calibration, supported by robust cross-validation and temporal validation results. The model identified five major predictors-female sex, higher ASA classification, elevated preoperative lactate, prolonged operative duration, and greater frailty (mFI ≥0.25), which jointly delineated patients at high risk of poor recovery. Beyond strong predictive performance, this study highlights the feasibility of using interpretable ML methods to deliver clinically actionable insights that extend beyond traditional risk models.

Our findings reaffirm that commonly available perioperative variables contain substantial prognostic information regarding postoperative recovery quality. Female sex has been repeatedly linked to worse cardiac surgical outcomes, possibly due to smaller vessel size, older age at surgery, and higher comorbidity burden. More and more evidence^15–18^ indicates that the prognosis of cardiac surgery in women is worse than that in men. Women often undergo surgery at older age and with more comorbidities, and sex-specific anatomical differences have been linked to less favorable outcomes. Similarly, the ASA class predicted recovery quality as expected: ASA III-IV patients carry greater disease burden, and classic studies have demonstrated that each step increase in ASA multiplies the risk of postoperative complications^19^. Likewise, elevated preoperative lactate likely reflects tissue hypoperfusion or metabolic stress. Prior work found that even baseline arterial lactate is an independent predictor of early adverse outcomes after cardiac surgery^20^, implying that patients entering surgery with higher lactate are already physiologically strained. High frailty index scores signify limited physiologic reserve, which we found markedly worsened recovery^8, 21^. This aligns with recent reports in cardiac and critical-care cohorts: frailty (as measured by mFI) is strongly associated with complications and death^22^. A retrospective cohort study observed that each one-point increase in mFI raised in-hospital mortality odds by 13% in acute myocardial infarction patients^23^. Finally, longer surgical duration was linked to poorer QoR, likely due to the cumulative burden of prolonged anesthesia, blood loss, and systemic inflammation^24^. A meta-analysis^25^ confirmed a robust relationship between operative time and complications: the odds of complications roughly double when surgery exceeds two hours, and increase 14% for every additional 30 minutes. Taken together, these interpretations indicate our key predictors have plausible mechanistic connections to recovery and corroborate existing knowledge.

Collectively, these predictors reflect the interaction between baseline physiological vulnerability (ASA class, frailty, sex) and intraoperative stressors (lactate, duration). Their convergence supports the biological plausibility and interpretability of the model’s outputs, a critical requirement for clinical adoption.

Traditional risk assessment tools^26^ such as the EuroSCORE II and STS models primarily target mortality or major adverse events, offering limited insight into patient-centered recovery outcomes^27^. In contrast, our model focuses on the multidimensional concept of recovery quality, capturing physical, emotional, and functional domains via the QoR-15 scale. Prior studies^28^ have attempted to apply regression-based nomograms to predict QoR-15 outcomes, but these methods often overlook nonlinear relationships and feature interactions. Our ML model builds on this by incorporating nonlinear interactions and achieving higher predictive accuracy. In general, these comparisons show our findings are in line with the literature, and our work uniquely extends ML prognostics to the domain of postoperative recovery in cardiovascular surgery.

The implications of these findings are both theoretical and practical. Academically, the study demonstrates that machine learning techniques, coupled with SHAP explainability, can elucidate the relative importance of perioperative risk factors in shaping recovery.

The translational potential of our findings lies in two domains: individualized risk stratification and perioperative decision support. First, the model’s outputs can identify patients at elevated risk for poor recovery prior to or immediately after surgery. For example, a frail female patient with elevated lactate could be flagged for enhanced postoperative surveillance, early mobilization, or nutritional and rehabilitation interventions. Such targeted resource allocation aligns with the principles of Enhanced Recovery After Surgery (ERAS) and value-based perioperative care.

Second, the model’s interpretable framework enables real-time clinical deployment. Integration into an electronic health record (EHR) system could facilitate automatic computation of recovery risk immediately upon completion of perioperative data entry. Importantly, the SHAP-based visualizations allow anesthesiologists, surgeons, and nurses to identify modifiable risk factors, such as optimizing metabolic status, shortening operative duration when feasible, or implementing prehabilitation strategies for frail patients. In this way, the model not only predicts outcomes but also informs actionable interventions, bridging predictive analytics and clinical decision-making.

Despite achieving exceptionally high discrimination (AUC 0.982), we recognize the potential risk of model overfitting. To mitigate this, we employed stratified sampling, 10-fold cross-validation, and permutation testing to confirm stability, with performance differences <0.02 between folds and validation sets. The model also underwent temporal external validation, demonstrating consistent performance across later patient cohorts.

However, the validation cohort was derived from the same institution and thus represents temporal rather than geographical generalizability. Further multicenter studies involving diverse patient populations are warranted to confirm external robustness and reproducibility. The consistency of predictor importance across time supports internal validity, but generalization to different clinical environments should be empirically confirmed before clinical deployment.

Several limitations should be noted. First, the retrospective single-center design may introduce selection bias and restrict generalizability. Second, some potentially relevant factors such as intraoperative hemodynamic trends, anesthesia techniques, or surgeon experience were not captured in the current dataset. Inclusion of high-frequency intraoperative monitoring data in future models could improve accuracy and dynamic prediction capabilities. Third, QoR-15 was assessed at a single postoperative time point (day 3), which reflects early recovery but not long-term quality-of-life outcomes. Finally, although model interpretability was enhanced through SHAP, this approach assumes additive feature effects and may not fully capture higher-order interactions.

Future work will focus on prospective, multicenter validation to establish external generalizability and evaluate real-world clinical impact. Integrating the model into perioperative information systems could enable automated risk alerts and personalized intervention protocols. Moreover, combining structured EMR data with intraoperative waveform and biomarker data may allow real-time dynamic prediction of recovery trajectories. Prospective interventional studies should also test whether modifying identified risk factors such as preoperative frailty reduction or lactate optimization-translates to improved QoR-15 outcomes. Such translational research will be essential to bridge algorithmic prediction and outcome improvement.

### Conclusions

In summary, this study demonstrates that an interpretable XGBoost model can accurately predict early postoperative recovery quality after cardiovascular surgery using routinely collected clinical data. The model identifies key physiological and procedural determinants of recovery and provides an interpretable, data-driven framework to guide individualized perioperative care. With further external validation and clinical integration, such predictive tools have the potential to enhance recovery-oriented outcomes and optimize perioperative resource utilization.

## Acknowledgements

We would like to express our sincere gratitude to the Cardiac surgery team of the Affiliated Hospital of Yangzhou University. At the same time, We sincerely thank all the patients.

## Author contributions

Luo Zhang conceptualized the study, developed the methodology, applied the software, performed the statistical analysis, visualized the data, wrote the original draft. Bei Ma and Zhi Xing collected the data and performed the statistical analysis. Yudong Wang and Shunping Tian developed the methodology and performed the statistical analysis. Zhuan Zhang conceptualized the study, reviewed and edited the manuscript. Jianyou Zhang confirm the authenticity of all the raw data. All authors have read and approved the f inal version of the manuscript.

## Data availability

All datas in this study should be requested from the corresponding authors.

## Code availability

All custom codes in this study should be requested from the corresponding authors.

## Declarations

### Ethics approval and consent to participate

The study was reviewed and approved by the institutional Ethics Committee(No.2023-YKL01-09), and conducted in accordance with the Declaration of Helsinki. The ethics committee waived the requirements for informed consent and clinical trial registration due to the retrospective nature of the study.

### Consent for publication

All authors have approved the manuscript for submission.

